# Longitudinal changes in network-based functional connectivity over a rugby season in adolescent males

**DOI:** 10.1101/2025.03.16.25324069

**Authors:** Joshua P. McGeown, Mangor Pedersen, Melanie Bussey, William S. Schierding, Paul Condron, Taylor Emsden, Tuterangi Nepe-Apatu, Maryam Tayebi, Stephen Kara, Matthew A. McDonald, Vickie Shim, Samantha J. Holdsworth, Eryn E. Kwon

**Author notes:** Corresponding author – Joshua P. McGeown, Address – Mātai Medical Research Institute, 466 Childers Road, Gisborne New Zealand, 4010 Email –.

## Abstract

**Objectives:** This study examined the impact of a single season of rugby union – and subsequent exposure to head acceleration events (HAEs) – on functional connectivity in adolescent males compared to non-collision sport athletes.

**Methods:** Resting-state functional MRI scans were acquired from 72 rugby players and 20 non-collision sport athletes. The rugby cohort were scanned longitudinally throughout the season. Voxel-wise maps of functional connectivity (FC) for 15 resting-state brain networks were generated for 178 datasets. Cross-sectional comparisons were performed between the rugby cohort at different stages of the season and the non-collision sport group. Longitudinal analyses were performed within the rugby cohort. An exposure analysis estimated HAE exposure based on the number of matches played.

**Results:** No significant cross-sectional differences in FC were found between rugby and non-collision sport athletes or between rugby players with high versus low exposure to HAEs. Longitudinally, rugby players showed increased inter-network FC over the season, with strengthening of connectivity in the temporal, motor, secondary visual, and anterior intraparietal sulcus networks, and mid-season decreases in the cerebellar-visual network. No association was found between longitudinal FC changes and changes in self-reported symptoms.

**Conclusions:** These findings suggest that participation in a season of rugby is associated with neuroplastic changes. These changes may reflect compensatory adaptations to preserve neurological function during periods of HAE exposure. Alternatively, they may also represent developmental changes or responses to physical activity and/or motor learning. This highlights the complexity of interpreting changes in FC in adolescent athletes participating in collision sports and the need for further research.

## Introduction

Collision sports are popular forms of recreation and involve purposeful and routine collisions between athletes and/or inanimate objects as part of normal gameplay and tactics [1]. Rugby codes, American Football, football (soccer), and ice hockey are popular forms of collision sports, with a combined estimate of 276 million participants globally [2–6]. Regular participation in sport and/or physical activity is associated with improved cardiovascular and metabolic health and reduced smoking and drug use [7, 8]. However, concerns have been raised that participation in collision sports is detrimental to neurological health [9–13].

Collision sports are associated with repetitive exposure to head acceleration events (HAEs), defined as short-duration events in which the head is accelerated either by a direct external force to the head or indirectly through force applied to the body [14]. The magnitude and direction of a given HAE imposes varying degrees of strain on neural tissue. A single high-magnitude HAE may result in clinically diagnosed mild traumatic brain injury (mTBI; [15]), which occurs at higher rates in collision sports compared to non-contact sports such as swimming, cross-country running, or volleyball [16, 17]. However, most collision sport-related HAEs are “subclinical” where the force transferred to the brain is insufficient to produce noteworthy signs and symptoms of a clinically meaningful injury [18, 19]. Evidence suggests that cumulative exposure to HAEs may contribute to negative cognitive outcomes and higher rates of neurodegenerative disease [10–13]. Reports from retrospective case-series studies indicates that exposure to HAE contributes to pathological changes in the brain [20–23]. Some evidence exists that the degree of pathology is more closely associated with greater cumulative exposure to HAE (i.e. number of seasons of participation in collision sports) than to the number of mTBIs experienced [20]. While the strength of these associations is unclear [24], the literature highlights a need for prospective studies to investigate how HAE exposure affects brain health in collision sport athletes over time.

Advances in neuroimaging, particularly magnetic resonance imaging (MRI), are increasingly enabling the non-invasive study of mTBI and HAE effects on brain structure and function *in vivo*. While conventional clinical MRI techniques are often insufficient to detect the subtle effects of mild brain trauma [25, 26], advanced MRI methods hold significant promise for this purpose [27–29]. One such technique is resting-state functional MRI (rs-fMRI), which measures spontaneous blood-oxygen-level-dependent (BOLD) signals when the brain is in a ‘resting’ or task-free state. These BOLD signals can be used to investigate how participation in collision sport and exposure to HAEs may affect functional connectivity within and between brain regions or across functionally related brain networks.

In New Zealand, rugby union is a popular collision sport and is considered the national game, with 155,000 participants in a country of just 5 million people [30]. This study examined how participation in a single rugby union season affects functional connectivity in comparison to a group of non-collision sport athletes. These findings represent early results from an ongoing longitudinal cohort study investigating the relationship between participation in collision versus non-collision sports and brain health in adolescent athletes. We focused on adolescent males, who represent 20% of participants in New Zealand, as it is crucial to determine whether participation in collision sports, and the associated exposure to HAEs, negatively impacts the developing brain. We used the number of contact trainings and matches as a proxy for HAE exposure over the season, given evidence linking greater exposure to worse long-term outcomes [20].

Based on previous research demonstrating altered connectivity following HAE exposure [31–34], we hypothesised that HAE exposure disrupts the functional organization of the brain which would be represented by decreased network-based functional connectivity in cross-sectional comparisons (collision sport athletes relative non-collision sport athletes) and longitudinal analysis. Additionally, we predicted that reduced functional organization would correlate with greater self-reported symptoms commonly associated with mTBI [31].

## Methods

This work was conducted under ethical approval from the New Zealand Health and Disability Ethics Committee (Ethics Ref: 20/NTB/14/AM09) in accordance with the Declaration of Helsinki. Participants aged ≥16 years provided written informed consent, while those <16 years provided written assent with parental consent.

### Design and participants

Seventy-two adolescent male rugby union athletes were recruited across two competitive secondary school seasons (2021 and 2023). Hereafter, this group will be referred to as the collision sport cohort. Twenty age-, sex-, and ethnicity-matched athletes engaged in non-collision sports were recruited as a comparison group. Potential participants were ineligible for this study if they had a pre-existing diagnosed neuropsychiatric condition and/or if they had dental braces that would cause imaging artifacts. Potential participants were excluded from the collision sport cohort if they self-reported an mTBI within six months of study initiation, while potential participants for the comparison group were excluded if they self-reported an mTBI or participation in a collision/martial sport within three years of joining the study. The comparison group were actively participating in non-collision sports such as basketball, field hockey, rowing, swimming, and volleyball. No football (soccer) athletes were included in the comparison group due to potential confounding effects of heading.

The collision sport cohort completed multimodal MRI acquisition sessions and questionnaires to assess symptoms commonly associated with mTBI in the early-season and post-season. Each session required approximately one hour, with 45 minutes allocated to image acquisition. The term *early-season* is used rather than *pre-season* because a larger group of players participated in a training camp that involved full collision training to determine which players made the teams that were a part of the study. Logistical constraints meant baseline assessments took place after team selection but before the first competitive match. A subset of the collision sport cohort completed an additional MRI and symptom assessment session during a brief mid-season break (Figure 1). The comparison group completed a single MRI acquisition session. All demographic and symptom data were collected and managed using REDCap electronic data capture tools hosted at Yale University [35]. A research team member was embedded with the collision sport cohort to record field notes (*i.e.* attendance) at all collision training sessions and matches.

**Figure 1.**
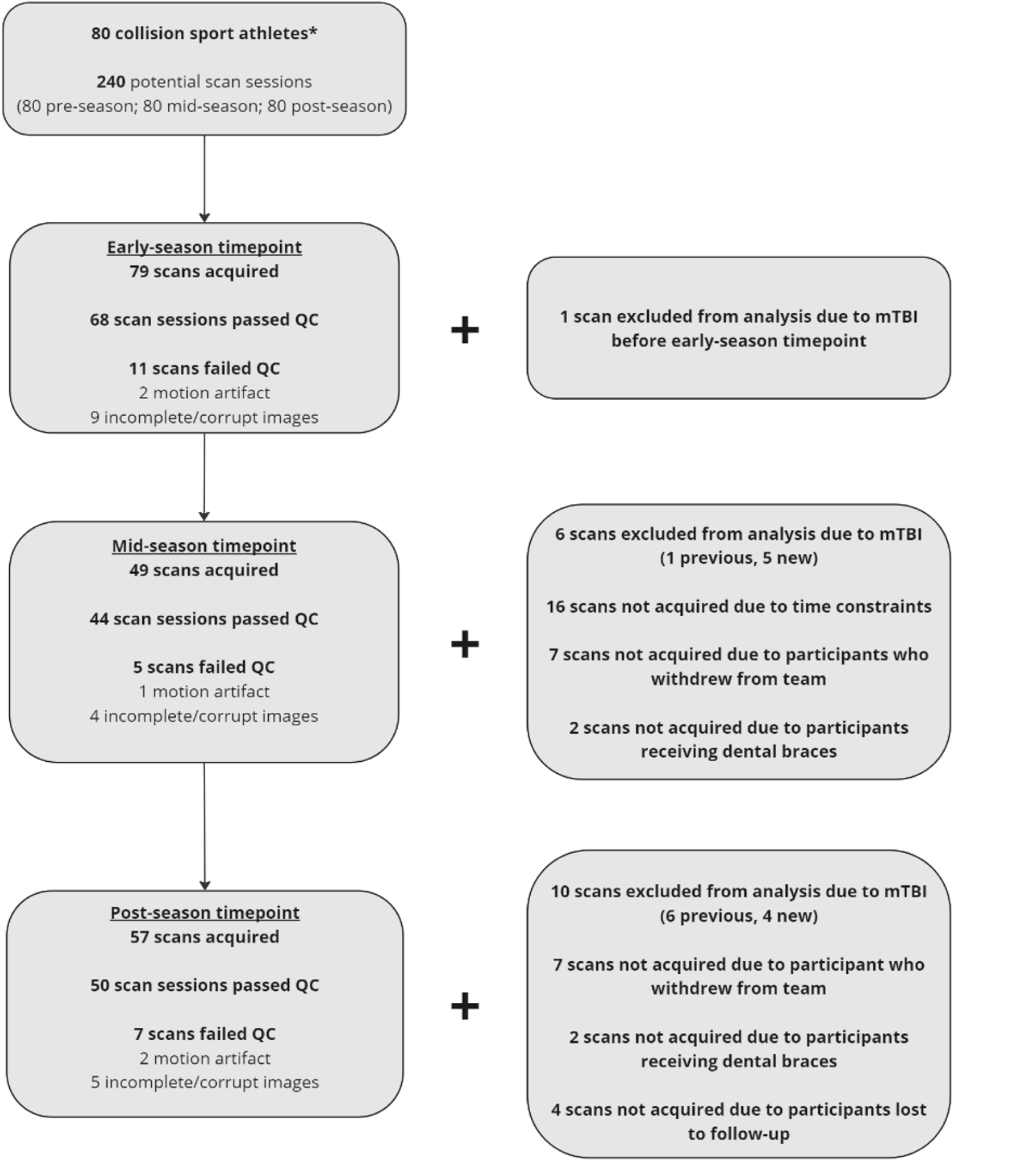
Flowchart of data availability and quality control.

### Imaging protocol and behavioural assessment

All MRI scans were performed on a single 3.0T GE Signa Premier MRI scanner (General Electric, MI, USA) using an AIR™ 48-channel head coil. An Axial Resting State (2D EPI GRE rs-fmRI) sequence was acquired with the following parameters: Repetition Time (TR) of 1529 ms, Echo Time (TE) of 30 ms, flip angle of 80°, matrix size of 64 × 64, Field of View (FOV) of 240 mm × 240 mm, number of slices 50-52, voxel size of 3.8 mm × 3.8 mm × 3 mm, and a HyperBand (simultaneous multi-slice) factor of 2. The scan time for the rs-fMRI sequence was 5:12 min and included four dummy samples and 200 statistical samples. Additionally, a 3D Axial T1 Bravo (IR-prep fast SPGR) sequence was acquired with TR of 6.8 ms, TE of 2.8 ms, flip angle of 12°, Inversion Time (TI) of 600 ms, matrix size of 256 × 256 (Zip512), FOV of 230 mm × 200 mm, 320 locs per slab, voxel size of 0.9 mm × 0.9 mm × 1 mm, parallel imaging factor (ARC) = 2, and number of excitations (NEX) = 1, with a scan time of 3:42 min. Additional clinical images were acquired and evaluated for incidental findings by a board-certified radiologist. No clinically relevant incidental findings were observed in the collision sport cohort or the comparison group.

Symptoms commonly associated with mTBI were assessed at each timepoint using the Sport Concussion Assessment Tool (SCAT5) Symptom Scale [36]. Symptom burden was determined using the total number of symptoms reported (out of 22 possible symptoms) and total symptom severity (out of a maximum severity score of 132).

### Functional MRI preprocessing and connectivity analysis

All preprocessing and first-level analyses were performed using our restingSnake workflow (https://github.com/MataiMRI/restingSnake.git). restingSnake leverages the Python-based Snakemake [37] workflow management system to enable reproducible and scalable preprocessing of rs-fMRI datasets using open-source neuroimaging tools. Anatomical and functional images were converted from dicom series to Nifti format and organised based on the Brain Imaging Data Structure [38] using HeuDiConv software [39]. Quality of each T1w and BOLD dataset was assessed based on visual reports generated by MRIQC [40]. Reports were visually inspected to confirm T1w alignment and to rule out signal artifacts. Carpet plots for BOLD timeseries were evaluated to assess motion-based quality control measures. Participant data were excluded if any BOLD timepoint exceeded 5mm of framewise displacement or if the mean framewise displacement was greater than 0.5mm.

Each pair of T1w and BOLD images that passed quality control were preprocessed using the fMRIPrep v23.1.3 pipeline [41]. Briefly, each T1w image was intensity corrected and skull-stripped. The brain-extracted T1w images were then used for tissue segmentation and surface reconstruction. Each T1w image was spatially normalised to MNI space using non-linear registration to the MNI152NLin2009cAsym template. Head motion parameters were estimated for each BOLD image, and the slice-timing correction was applied. The slice-timing corrected BOLD image was resampled to native space and then co-registered to the T1w reference using boundary-based registration. Motion-, tissue-, and component-based confounding timeseries were estimated for each BOLD image. Finally, the preprocessed BOLD timeseries was resampled into MNI152NLin2009cAsym space.

Before first-level analysis, visual reports generated by fMRIPrep were inspected to ensure correct registration, tissue segmentation, and surface labelling. No datasets were excluded at this stage. First-level analysis, including denoising, was implemented using the Nilearn Python package (https://github.com/nilearn/nilearn.git). For first-level analysis, the nilearn.maskers module was applied to each BOLD timeseries. Masker objects were configured with a 6 mm full-width-half-maximum smoothing kernel, and included steps for detrending, standardization, and bandpass filtering (0.01 to 0.1 Hz) the BOLD signal.

Confound timeseries, including white matter and cerebrospinal fluid signals, as well as six motion parameters (x, y, z translations and rotations), were regressed from the BOLD signal. The residual BOLD signal was then used for network-based functional connectivity analysis. This analysis utilised the Multi-subject Dictionary Learning (MSDL) atlas [42] to balance the trade-offs between data-driven identification of resting-state functional brain networks and reproducibility across datasets. *Figure 2* visualises the 15 resting-state networks within the MSDL atlas that were analysed in the present study.

**Figure 2.**
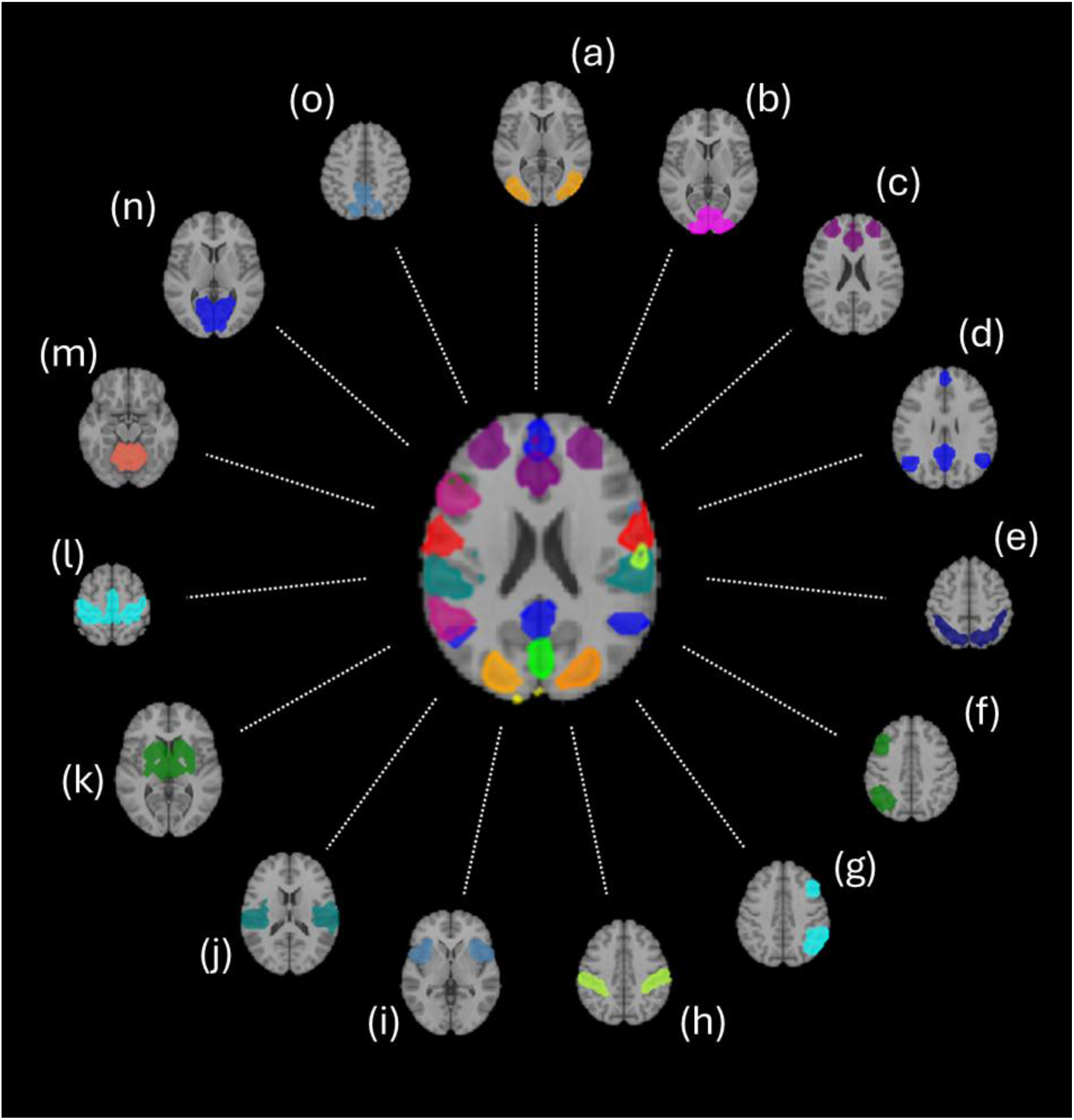
Functional connectivity networks-of-interest from MSDL atlas. a) Secondary Visual Network; b) Posterior Occipital Network; c) Salience Network; d) Default Mode Network; e) Dorsal Attention Network; f) Left Ventral Attention Network; g) Right Ventral Attention Network; h) Anterior Intraparietal Sulcus Network; i) Cingulate-Insula Network; j) Temporal Network; k) Basal Ganglia Network; l) Motor Network; m) Cerebellar Network; n) Striate Network; o) Dorsal Posterior Cingulate Cortex Network.

The BOLD timeseries for each node of the networks were extracted using nilearn.maskers.NiftiMapsMasker, resulting in a numpy array with dimensions (n_volumes, n_nodes). Similarly, nilearn.maskers. NiftiMasker produced a brain-wide voxel-wise timeseries array with dimensions (n_volumes, n_voxels). Node-to-voxel correlation analysis was performed using these arrays to produce a correlation matrix with dimensions (n_voxels, n_nodes), representing correlation values between each network node and each voxel’s signal, ranging from –1 to 1. Fisher’s transformation was applied to convert the correlation matrices to *z*-maps. These z-maps were then averaged across the nodes of each network, resulting in a single network-based brain-wide voxel-wise *z*-map.

### Statistical analysis

Eight athletes in the collision sport cohort (n = 72) participated in the 2021 and 2023 data collection seasons. Data from these participants were included from both seasons and analysed as distinct datasets, meaning a total potential dataset of 240 unique sessions (80 participants across three timepoints) for the collision sport cohort. Figure 1 depicts the sample sizes available for each group-level voxel-wise analysis performed using the FMRIB Software Library (FSL). Mass univariate permutation testing was conducted using FSL’s randomise, with 5000 permutations and threshold-free cluster enhancement (TFCE). A conservative threshold of p < 0.01 was applied to TFCE corrected maps to identify significant clusters and control for Type I error. Figure *3* summarises the preprocessing and statistical analysis workflow. Two– and three-group comparisons of continuous demographic variables were performed using Mann-Whitney U and Kruskal-Wallis tests, respectively. Distributions of categorical variables were assessed with Chi-Squared tests.

**Figure 3.**
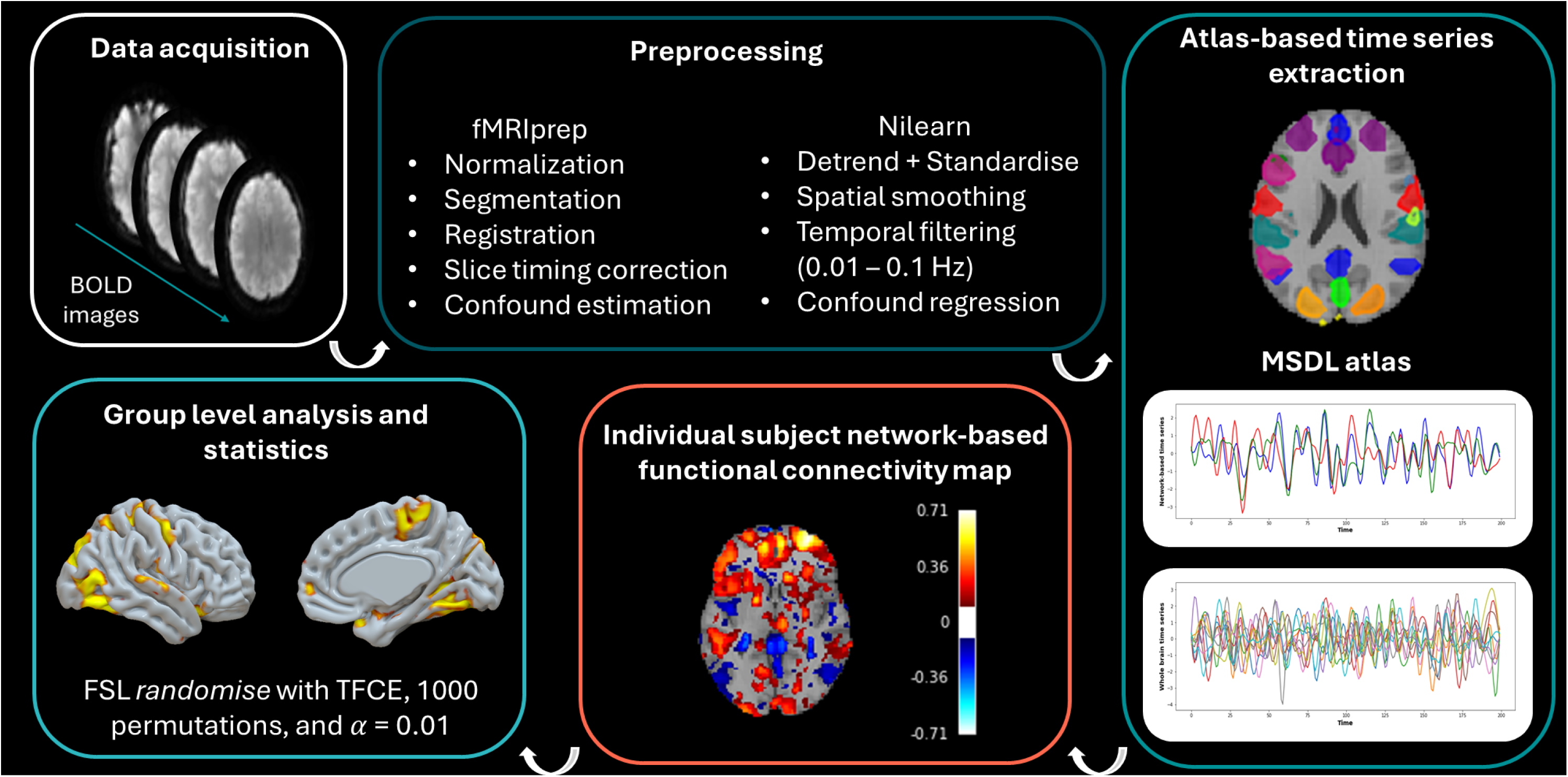
Data preprocessing and statistical analysis workflow.

Cross-Sectional Analysis: Pairwise two-sample t-tests were used to examine differences in network-based functional connectivity between the collision sport cohort and the comparison group at each timepoint: early-season (n = 68) vs. non-collision sport (n = 16), mid-season (n = 44) vs. non-collision sport, and post-season (n = 50) vs. non-collision sport.

Longitudinal Analysis: One-sample t-tests assessed changes within the collision sport cohort across the season. This involved comparing difference maps for: mid-season vs. early-season (paired n = 42), post-season vs. mid-season (paired n = 35), and post-season vs. early-season (paired n = 47).

Exposure Analysis: A one-way ANOVA with three groups was conducted to determine if post-season connectivity differences relative to the comparison group were mediated by exposure to HAEs during the season. Participants were categorised into high-(n = 25) and low-exposure (n = 22) subgroups based on whether they played above or below the median number of matches. Match participation was considered a binary variable where any time spent on the field during live gameplay for a given match was considered as a “match played”. Fractions of matches played were not considered in the current analysis.

Effect Sizes and Correlations: Cohen’s D effect sizes were calculated for significant clusters. To evaluate potential clinical implications, Spearman rho correlations were also used to examine the relationship between changes in network-based functional connectivity and SCAT5 symptom scores.

## Results

### Demographic comparisons

There was no significant difference for ethnicity or number of self-reported mTBI between the collision sport cohort and comparison group (Table *1*; *p >* 0.05). On average, the comparison group was slightly older (Median: 17 years, IQR [16,18]) than the collision sport cohort (Median: 16 years, IQR [16,17]; *p* < 0.05), and subsequently the comparison group had one more year of education on average compared to the collision sport cohort (p < 0.05; Table *1*). The comparison group consisted of athletes participating in a range of non-collision sports including basketball, field hockey, rowing, swimming, and volleyball (Table *1*). There were no differences in framewise displacement between non-collision sport athletes (Median: 0.104, IQR [0.099, 0.128]) and the collision sport cohort (Median: 0.125; IQR [0.106, 0.157]; *p* > 0.05). There were also no differences in framewise displacement longitudinally for the collision sport cohort (Early-season Median: 0.127; IQR [0.106, 0.156]; Mid-season Median 0.121, IQR [0.096, 0.146]; Post-season Median 0.124, IQR [0.110, 0.172]; *p*’s > 0.05).

**Table 1.**
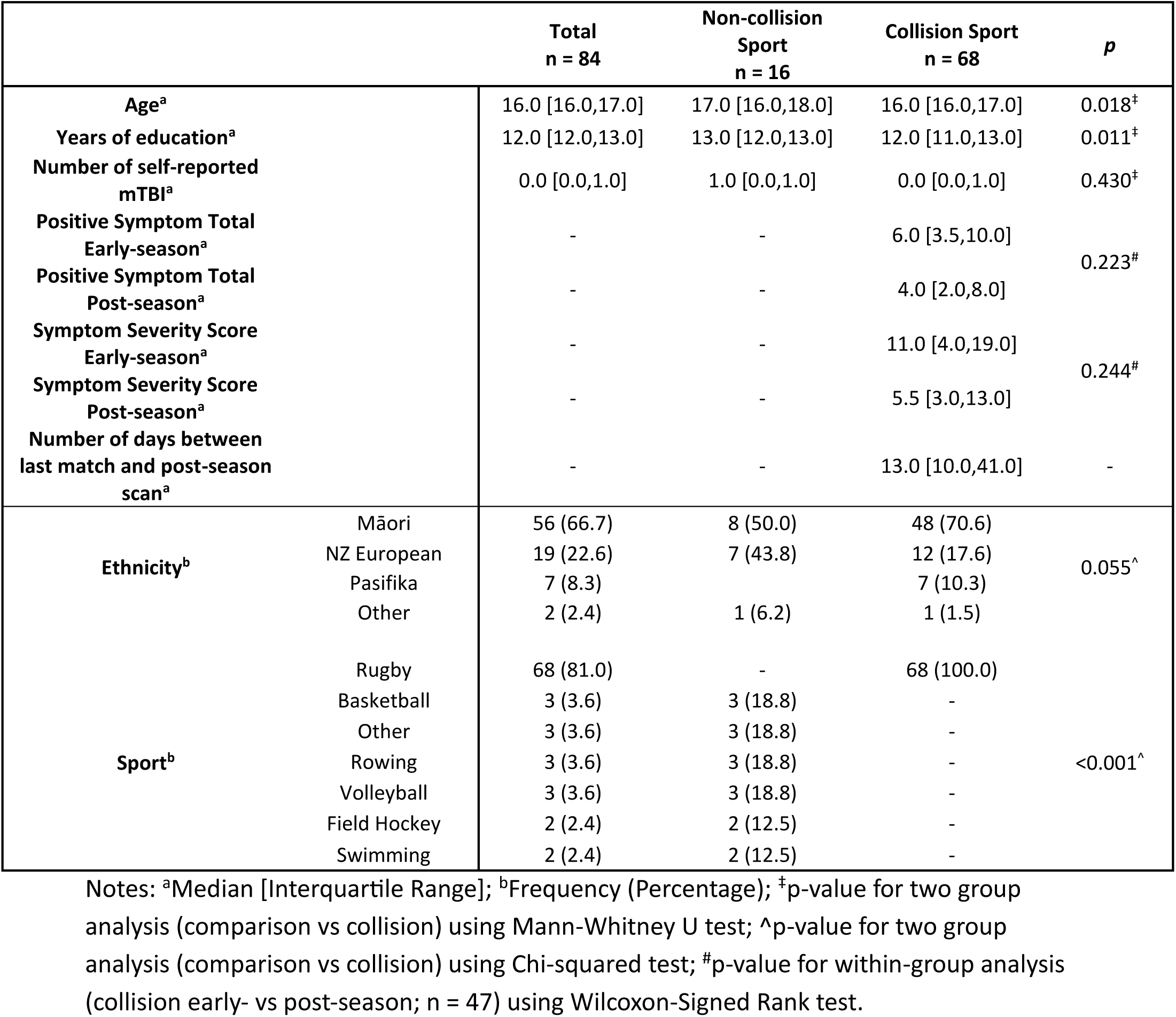
Sample characteristics.

### Cross-sectional functional connectivity analysis

No statistically significant differences in functional connectivity were observed for between-group pairwise analyses of the comparison group (n = 16) versus collision sport athletes at the beginning of the season (n = 64), mid-season (n = 44), or post-season (n = 50) for any of the 15 MSDL atlas networks.

### Exposure-based functional connectivity analysis

An exposure-based analysis was performed using post-season imaging data from 47 participants in the collision sports cohort. Exposure to HAEs was estimated based on the total number of matches played during the season. Visual inspection of the distribution of matches played revealed a clear bimodal distribution with a median of 9 (IQR: [6, 12]) matches for the whole sample. The high-exposure group was defined based on players who participated in ≥9 matches (Table *2*; n = 25; Median: 12; IQR [11, 13]) and the low-exposure group consisted of participants who played in <9 matches (n = 22; Median: 6; IQR [4, 7]). Estimating exposure using number of contact trainings attended or combined total of matches and contact trainings did not change subgroup stratification. There were no differences in ethnicity, years of education or self-reported history of mTBI between the comparison group, high-exposure, or low-exposure subgroups (Table *2*; *p*’s > 0.05). The comparison and high-exposure groups were on average older than the low-exposure groups (Table *2*; *p* < 0.05). The high-exposure group participated in significantly more matches than the low-exposure group (Table *2*; *p* < 0.001).

**Table 2.**
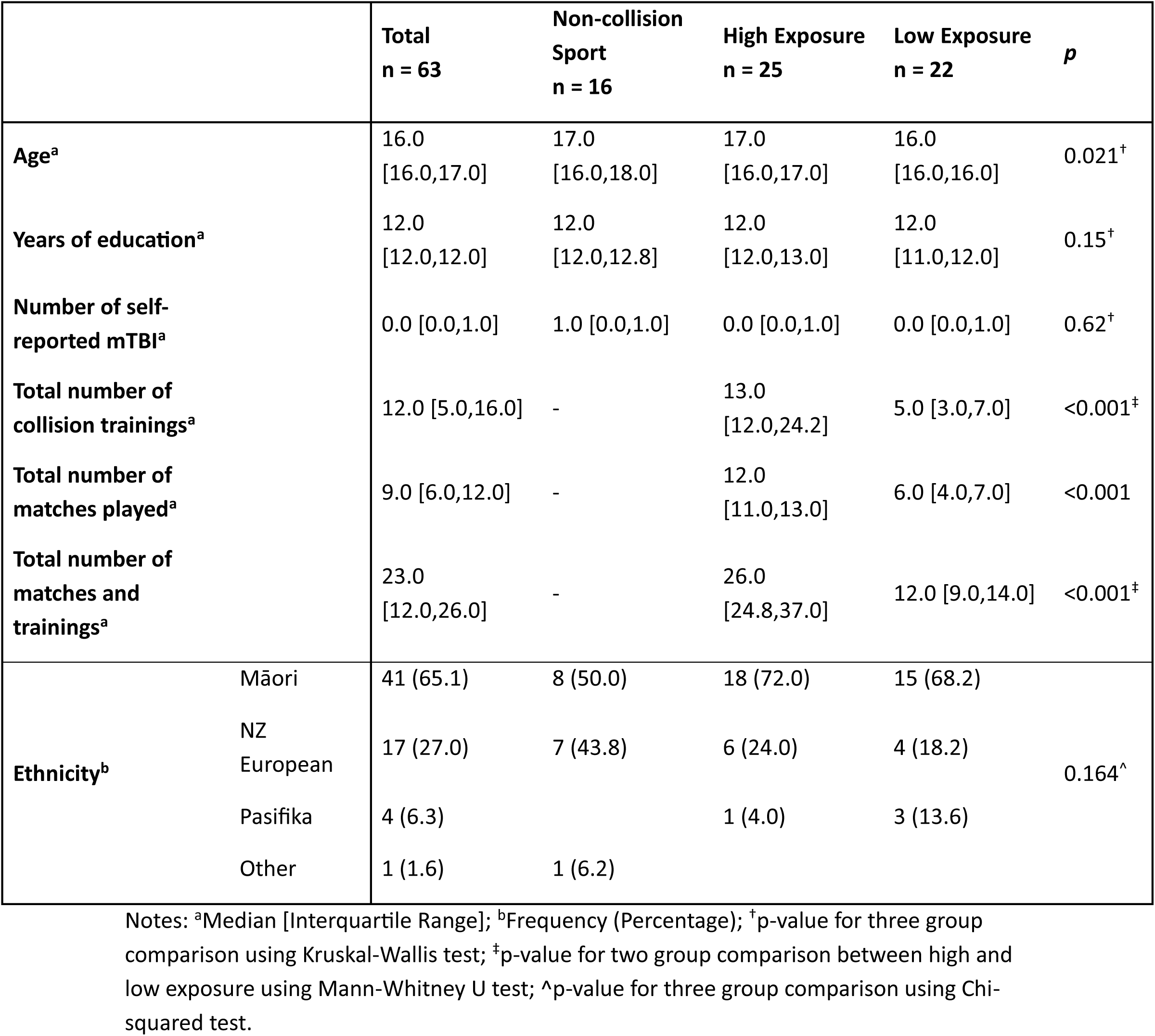
Sample characteristics based on exposure.

|  |  | **Total**<br>**n = 63** | **Non-collision**<br>**Sport**<br>**n = 16** | **High Exposure**<br>**n = 25** | **Low Exposure**<br>**n = 22** | **p** |
| --- | --- | --- | --- | --- | --- | --- |
| **Age<sup>a</sup>** |  | 16.0<br>[16.0,17.0] | 17.0<br>[16.0,18.0] | 17.0<br>[16.0,17.0] | 16.0<br>[16.0,16.0] | 0.021<sup>†</sup> |
| **Years of education<sup>a</sup>** |  | 12.0<br>[12.0,12.0] | 12.0<br>[12.0,12.8] | 12.0<br>[12.0,13.0] | 12.0<br>[11.0,12.0] | 0.15<sup>†</sup> |
| **Number of self-reported mTBI<sup>a</sup>** |  | 0.0 [0.0,1.0] | 1.0 [0.0,1.0] | 0.0 [0.0,1.0] | 0.0 [0.0,1.0] | 0.62<sup>†</sup> |
| **Total number of collision trainings<sup>a</sup>** |  | 12.0 [5.0,16.0] | - | 13.0<br>[12.0,24.2] | 5.0 [3.0,7.0] | <0.001<sup>‡</sup> |
| **Total number of matches played<sup>a</sup>** |  | 9.0 [6.0,12.0] | - | 12.0<br>[11.0,13.0] | 6.0 [4.0,7.0] | <0.001 |
| **Total number of matches and trainings<sup>a</sup>** |  | 23.0<br>[12.0,26.0] | - | 26.0<br>[24.8,37.0] | 12.0 [9.0,14.0] | <0.001<sup>‡</sup> |
| **Ethnicity<sup>b</sup>** | Māori | 41 (65.1) | 8 (50.0) | 18 (72.0) | 15 (68.2) | 0.164<sup>^</sup> |
|  | NZ European | 17 (27.0) | 7 (43.8) | 6 (24.0) | 4 (18.2) |  |
|  | Pasifika | 4 (6.3) |  | 1 (4.0) | 3 (13.6) |  |
|  | Other | 1 (1.6) | 1 (6.2) |  |  |  |

Notes: <sup>a</sup>Median [Interquartile Range]; <sup>b</sup>Frequency (Percentage); <sup>†</sup>p-value for three group comparison using Kruskal-Wallis test; <sup>‡</sup>p-value for two group comparison between high and low exposure using Mann-Whitney U test; <sup>^</sup>p-value for three group comparison using Chi-squared test.

There was no statistically significant effect of group membership based on exposure to HAEs on functional connectivity for any of the MSDL networks at the post-season timepoint. Post-season scans were acquired a median of 13 (IQR: [10, 41]) days after each participant’s final match of the season. A more conservative post-hoc analysis using only post-season imaging data acquired ≤21 days of the final match did not yield any different findings.

### Longitudinal functional connectivity analysis

Forty-two participants in the collision-sport cohort had longitudinal imaging data that passed quality control from early-to mid-season. Longitudinal analysis of these data identified a cluster of moderately reduced functional connectivity with the cerebellar network at mid-season relative to early-season (TFCE corrected, *p* < 0.01, *d* = –0.588; Figure *4*). Application of Harvard-Oxford Cortical labels [43] identified the left and right lateral occipital cortex, left and right occipital fusiform gyrus, left lingual gyrus, and right temporal occipital fusiform cortex were regions-of-interest exhibiting reduced functional connectivity with the cerebellar network (Table *3*). No other significant longitudinal network-based changes in functional connectivity were observed at mid-season relative to early-season.

**Figure 4.**
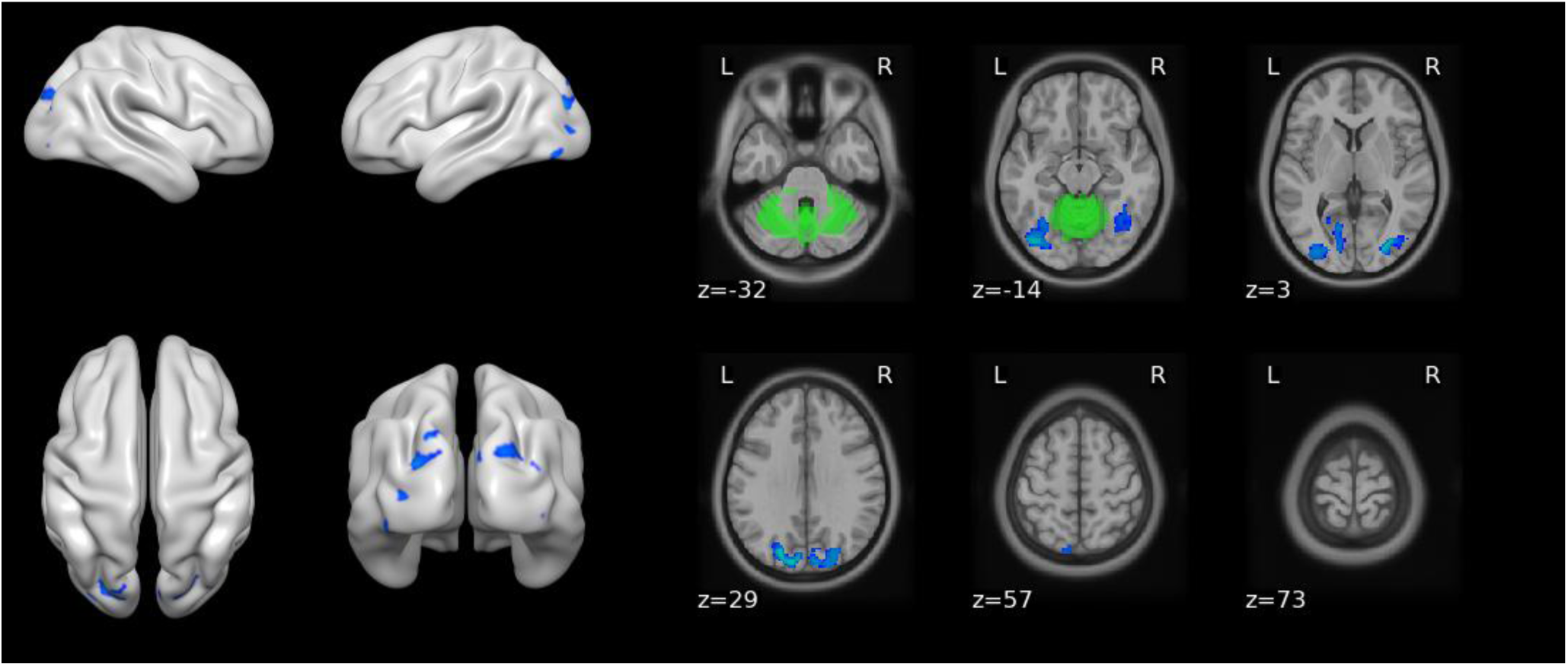
Regions with decreased connectivity (TFCE corrected, p<0.01) with cerebellar network at mid-season relative to early-season in collision-sport cohort. Green areas represent probabilistic representation of network according to MSDL atlas.

**Table 3.**
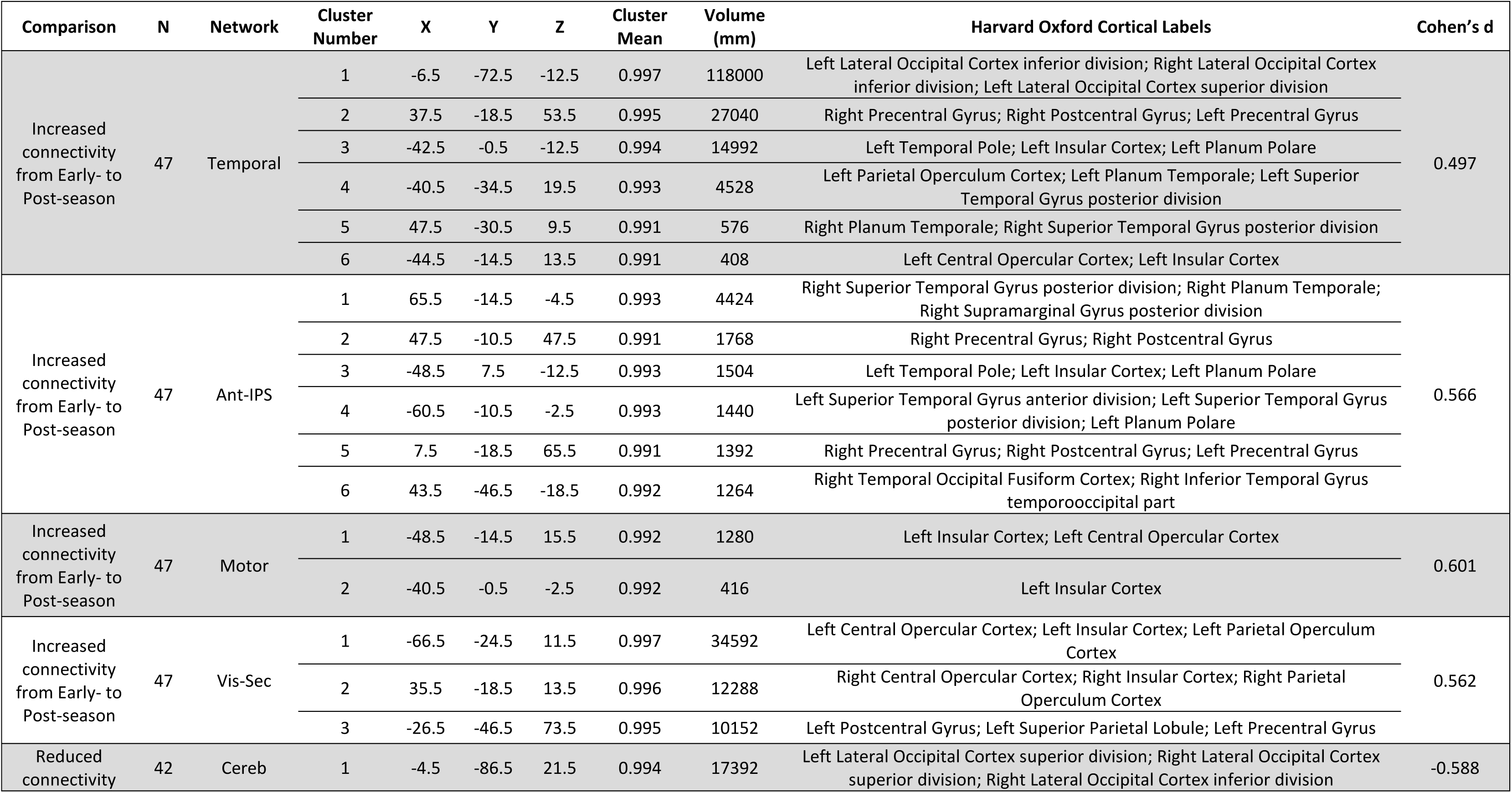
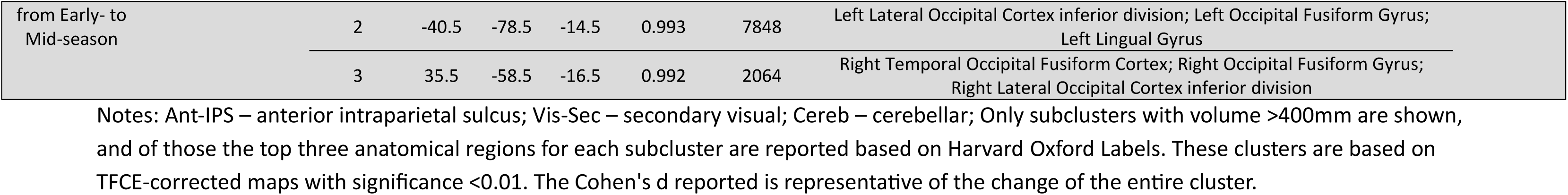
Anatomical labels for significant clusters in MNI space with effect sizes.

Forty-seven paired imaging datasets of sufficient quality were available from early-to post-season for the collision-sport cohort. No reductions in functional connectivity with any MSDL networks were evident at post-season relative to early-season. However, moderate increases in functional connectivity at post-season compared to early-season were observed in four clusters (TFCE corrected, *p*’s < 0.01) related to the anterior intraparietal sulcus network (*d* = 0.566; Figure *5*), motor network (*d* = 0.601; Figure *6*), secondary visual network (*d* = 0.562; Figure *7*), and temporal network (*d* = 0.497; Figure *8*) respectively. Increased connectivity with the anterior intraparietal sulcus network was pronounced bilaterally in the superior temporal gyrus, and the right pre/post-central gyrus (Table *3*). The left insular and central opercular cortex demonstrated increased connectivity with the motor network (Table *3*). Increased connectivity with the secondary visual network was evident bilaterally in the insular cortex, opercular cortex, and left pre/postcentral gyrus (Table *3*). Finally, increased connectivity with the temporal network was observed bilaterally in the lateral occipital cortex, planum temporale, superior temporal gyrus, and precentral gyrus as well as in the right postcentral gyrus, left insular cortex and left operculum cortex (Table *3*).

**Figure 5.**
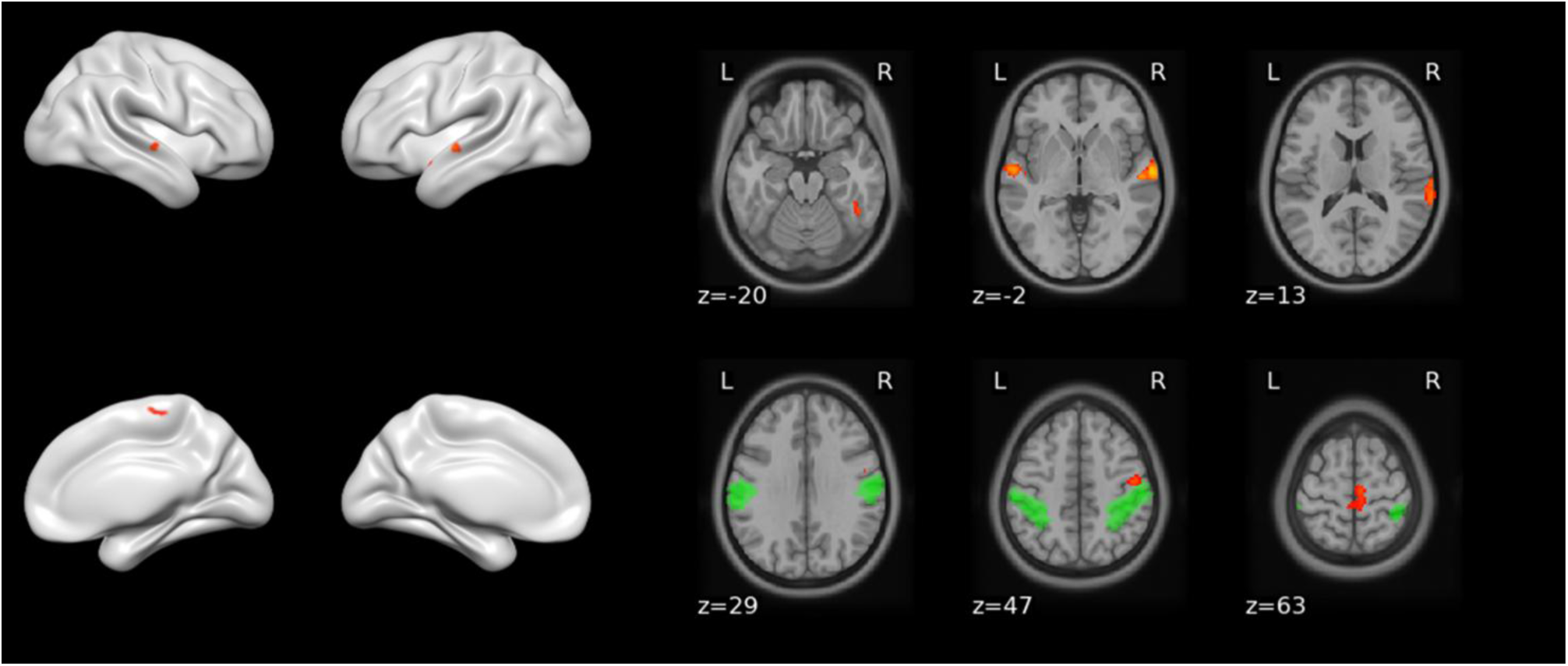
Regions with increased connectivity (TFCE corrected, p<0.01) with anterior intraparietal sulcus network at post-season relative to early-season in collision-sport cohort. Green areas represent probabilistic representation of network according to MSDL atlas.

**Figure 6.**
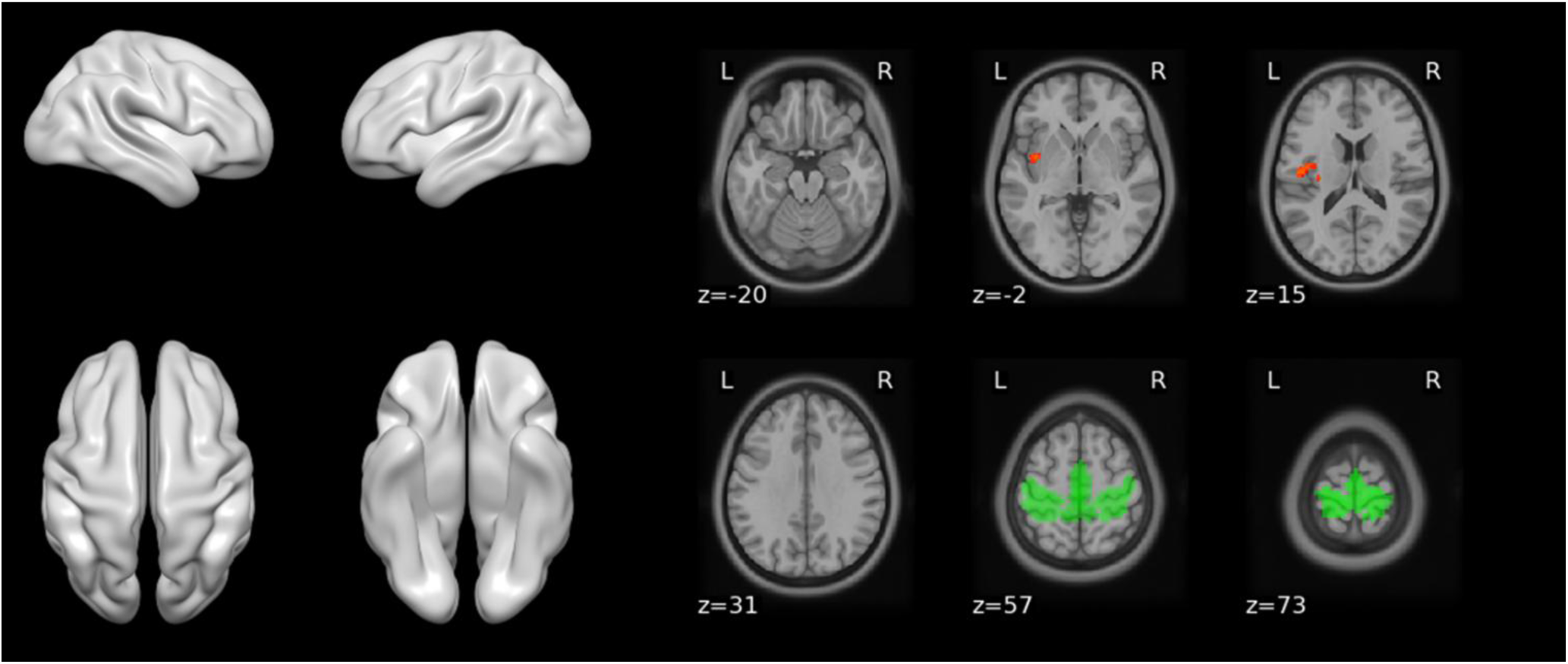
Regions with increased connectivity (TFCE corrected, p<0.01) with motor network at post-season relative to early-season in collision-sport cohort. Green areas represent probabilistic representation of network according to MSDL atlas.

**Figure 7.**
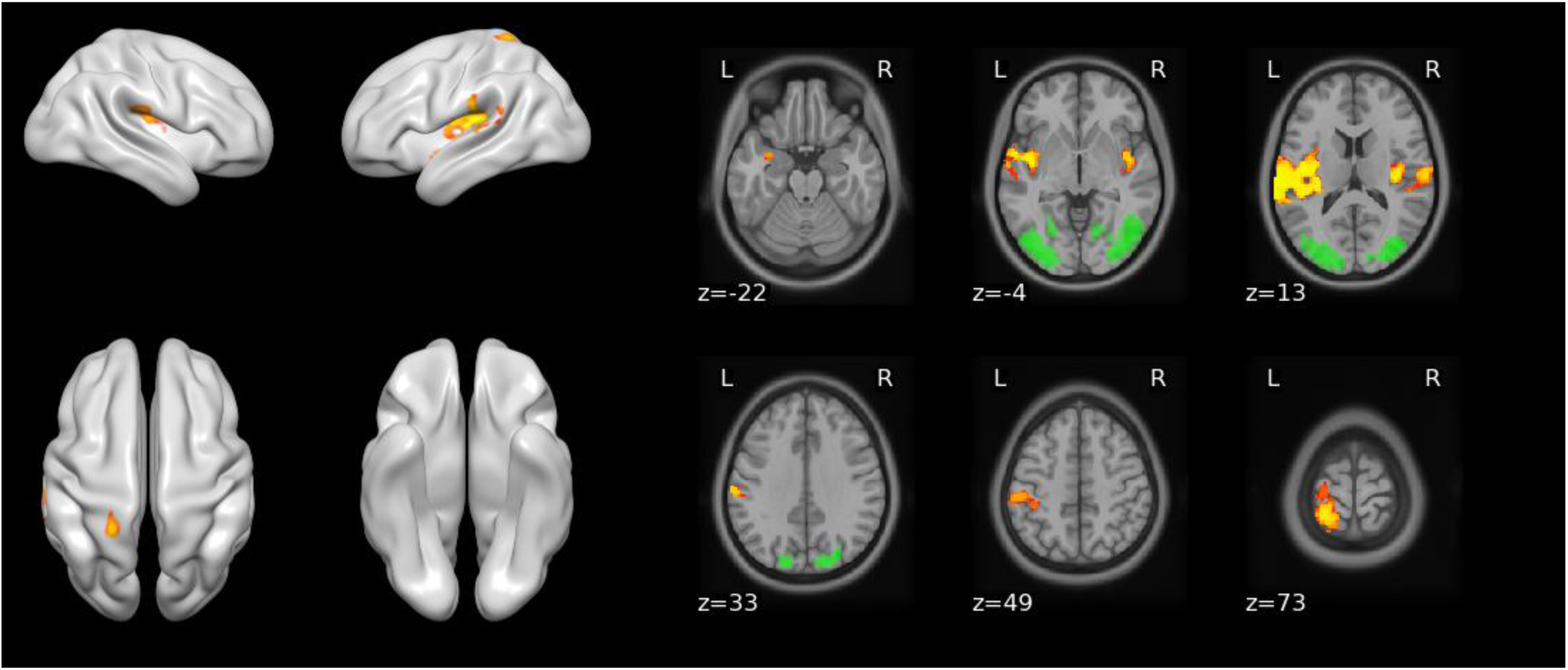
Regions with increased connectivity (TFCE corrected, p<0.01) with secondary visual network at post-season relative to early-season in collision-sport cohort. Green areas represent probabilistic representation of network according to MSDL atlas.

**Figure 8.**
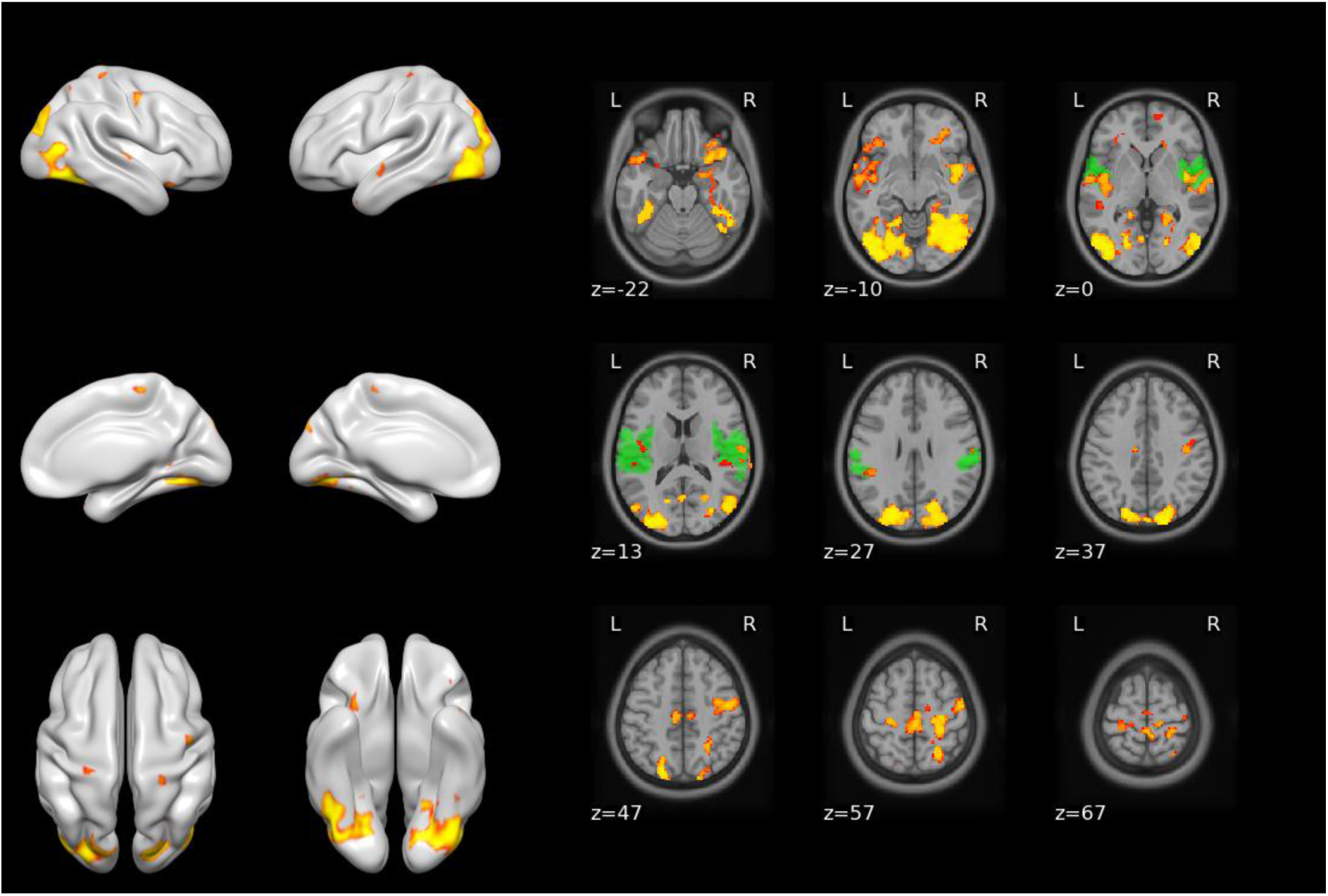
Regions with increased connectivity (TFCE corrected, p<0.01) with temporal network at post-season relative to early-season in collision-sport cohort. Green areas represent probabilistic representation of network according to MSDL atlas.

Longitudinal data were available for 35 participants in the collision-sport cohort from mid-season to post-season. No significant longitudinal differences were observed for any of the networks between these timepoints.

### Association between longitudinal changes and symptom load

A correlation analysis was performed to assess whether the observed longitudinal changes in functional connectivity were associated with changes in burden of common mTBI symptoms measured using the SCAT-5 Symptom Scale. Longitudinal imaging and symptom data were used from 47 collision-sport participants for this analysis. At a group level, there was no significant difference in the number of symptoms reported or the total severity of symptoms at post-season compared to early-season (*p* >0.05; Table *1*). Spearman’s correlations did not identify significant associations between change in subjective reports of symptom burden and longitudinal changes in functional connectivity (*p*’s > 0.05).

## Discussion

Use of neuroimaging to study the effects of collision sport-related HAE exposure on brain function is an emerging area of research. Our results demonstrate increased inter-network functional connectivity within rugby players at the conclusion of the season relative to the early season, mainly with bidirectional strengthening of the temporal, motor, secondary visual, and anterior intraparietal sulcus networks. Additionally, we observed reduced connectivity at mid-season relative to early-season between the cerebellar network and regions of visual networks. An investigation similar to ours that studied university-aged female rugby players found they exhibited greater connectivity between the default mode network and posterior cingulate cortex and between the lateral visual network and regions of the occipital lobe both during and after the season compared to non-contact athletes [44].

Interpretation of our findings in the context of previous work highlights three noteworthy discussion points regarding the complexity of studying the effects of collision sport participation on brain health in adolescents.

### Plausible explanations for functional hyperconnectivity in collision sport athletes

The biological explanation for longitudinal functional connectivity increases in rugby union athletes remains unknown. For example, Manning et al. proposed that hyperconnectivity may reflect compensatory mechanisms necessary to maintain normal neurological function during subclinical HAE exposure [44]. This theory was supported by a negative association between posterior cingulate cortex hyperconnectivity and reduced axial diffusivity within the splenium of the corpus callosum in the same subjects [44]. The authors suggested hyperconnectivity of widespread brain regions may be necessary to counteract HAE-related perturbations to axonal integrity to achieve normal neurocognitive function [44].

Hyperconnectivity is commonly observed following clinically diagnosed mild, moderate, and severe traumatic brain injury [27]. Studies have shown that, after mTBI, acute hyperconnectivity is associated with more favourable recovery outcomes [45, 46], whereas acute hypoconnectivity is linked to poorer outcomes [47–49]. These findings suggest periods of hyperconnectivity may be a desired homeostatic response to HAE exposure, whether subclinical or clinically apparent. Adaptive hyperconnectivity may explain our findings, with stronger cortical-cortical networks for sensorimotor integration and planning to compensate for hypoconnectivity between the cerebellar network and visual regions. Intepretation of the observed hyperconnectivity in collision sport athletes as a compensatory mechanism to counteract HAE exposure aligns with the connectomics of brain disorders described by Fornito et al. [50]. In their discussion, the authors outline how undamaged nodes within the injured system, or within neighboring systems, increase activity in order to preserve behaviour.

Alternatively, the observed cortical-cortical strengthening and cortical-subcortical weakening of functional connectivity in our cohort could represent typical development of brain networks during adolescence [51]. Additionally, we propose our results may be influenced by the profound effects regular participation in physical activity has on the brain [52, 53]. Brooks et al. found that regular physical activity and sports participation was associated with greater stability and efficiency of whole-brain and network-based functional connectivity in early adolescence [54].

Moreover, the longitudinal changes in our collision sport athletes may partly be explained by motor skill acquisition due to regular ball-handling and movement skills during trainings throughout the season. Several studies have demonstrated how skill acquisition training in football (soccer) players promotes notable functional connectivity increases within brain regions similar to our study [55–57]. These studies did not attribute changes in functional connectivity to potentially compensatory responses to HAE exposure during heading. This underscores how similar changes in functional connectivity observed in athletes who participate in collision sports can be interpreted differently depending on the research focus (*i.e.* HAE exposure vs motor skill acquisition).

Given the absence of cross-sectional differences, exposure-related differences, or negative associations between symptom reports and functional connectivity, we speculate one or more of these alternative contributing factors – typical development, physical activity, or skill acquisition – may explain our longitudinal results. While it remains premature to make definitive conclusions as to the degree to which these explanations account for our findings, the discussion above highlights that changes in functional connectivity may not be exclusively due to negative responses to HAE exposure.

### Number of matches as an estimate of HAE exposure

Our current exploration of potential exposure-mediated differences in functional connectivity was based on reports that the number of seasons, and thereby the number of matches, over a career of collision sport participation is an explanatory variable for post-mortem neurodegenerative pathology [20]. Our findings suggest that estimation of HAE exposure, based on number of matches played, is not associated with statistically meaningful differences in functional connectivity following a single season of rugby union relative to non-collision sport participation in adolescent males. This observation is not unexpected as the number of matches played is a crude approximation of HAE exposure. For instance, two players may both participate in seven matches, but one may spend 70 minutes on the field per match. At the same time, the other only plays 10 minutes each match for a total difference of 420 minutes of exposure time to potential HAEs between these hypothetical players. Person-time at risk is a key concept for injury rate and risk estimation in sports injury epidemiology [58]. In designing the exposure analysis of the current study, we followed the assumptions made by Mez et al. in their retrospective case-series, using the number of matches played as an estimate of HAE exposure, but this approach did not incorporate person-time at risk [20]. Furthermore, studies using instrumented mouthguards have highlighted significant variability in HAE incidence per player-hour in rugby union, particularly based on playing position [18], further underscoring the importance of accounting for individual differences in exposure.

While our current work does not capture these individual differences, it provides valuable insight by questioning the assumption that *any* participation in collision sports is inherently harmful [59]. Our findings suggest that, at least in adolescent male rugby union players, participation over a single season does not appear to be categorically associated with gross detrimental changes in functional connectivity. This interpretation is not intended to dismiss legitimate concerns about potential long-term consequences of HAE exposure [9–13]. Rather, it emphasises the need to better understand how exposure profiles may differentially impact brain health.

### Framing the issue of collision sport participation

Our results highlight the need to frame the topic of HAE exposure in collision sports in a wider context. Growing evidence indicating negative neurocognitive effects[10, 11] and an increased risk of neurodegenerative diseases later in life [12, 13] is the basis for concerns about the detrimental effects of subclinical HAE exposure in collision sports. These reports have prompted a rapid expansion of research efforts to understand the best strategies to reduce the risk of these negative outcomes. However, understanding the complex interplay between genetics, exposure, neurophysiology, and symptomology to explain why some athletes have unfavourable outcomes is still in the early stages. In the face of this uncertainty, some sporting organisations apply the precautionary principle by reducing or eliminating HAE exposure in youth sports including American Football, soccer, ice hockey, and rugby until these relationships are understood [60]. More extreme statements have come from some related interest groups, claiming collision sports constitute “child abuse” that can permanently injure children [59]. Such statements are an oversimplification of an inherently multivariate issue. While concerns about the neurological risks of HAE exposure are valid, it is equally important to recognise the broader scope of collision sport participation on health and well-being.

Several studies that highlight higher risk of neurodegenerative disease also report lower all-cause mortality and risk of heart or lung disease in these populations [13, 61]. Additionally, a recent systematic review concluded that sport of any form – including collision sports – benefit mental health and social functioning, raising the complex relationship between physical activity and overall well-being [62]. Multiple studies have reported progressive reduction in functional connectivity across several brain networks in American football players at later stages in the season relative to pre-season [31–34]. However, two of these studies observed patterns of functional connectivity indicative of recovery following the cessation of HAE exposure during the off-season [32, 33]. These observations of neurological recovery raise questions about the extent of irreversible harm associated with HAE exposure in collision sports.

Strauss et al. designed their neuroimaging study to frame the adverse effects of HAEs in the broader scope of sport participation [63]. They found that both non-collision sport and collision sport athletes with lower HAE exposure demonstrated a higher degree of white matter anisotropy and cognitive performance compared to a non-athletic comparison group.

Collision sport athletes with high HAE exposure did not differ from the comparison group regarding white matter microstructure or cognition [63]. These findings suggest the existence of a threshold below which the neurological benefits of physical activity may outweigh the risks of HAE exposure. Furthermore, their findings suggest that the benefits of physical activity may attenuate the detrimental effects of high levels of HAE exposure in a way that preserves neurocognitive function comparable to individuals who do not participate in sport.

Taken together, this summary of evidence highlights the complex continuum of risks and benefits associated with collision sport participation. Millions of individuals willingly elect to engage in these sports. Rather than perpetuating alarmist narratives, research efforts should focus on understanding mechanisms underlying subclinical HAE-related risks to inform evidence-based prevention and rehabilitation strategies. This would help maximise the benefits of participation while effectively managing and communicating the risks for those who engage in collision sports.

### Limitations and future directions

There are several limitations and opportunities for future research to recognise in the current study. First, HAE exposure was estimated based on the number of matches played by each player throughout the season. As discussed, this is an imprecise estimate of exposure compared to directly capturing HAE exposure using instrumented mouthguards. Kinematic mouthguard data have been collected from our cohort, and the association between these HAE metrics and imaging findings will be the focus of future works. Second, the sample size of our comparison group was relatively small compared to the collision sport cohort and may have contributed to the lack of significant cross-sectional findings. Additionally, the comparison group was only assessed at a single timepoint. Longitudinal data for the comparison group would greatly assist in the interpretation of the trends of hyperconnectivity observed in our collision sport cohort. Our group is recruiting more participants for the comparison group to be assessed longitudinally to address this limitation in future studies. Third, the proposed explanation that the observed hyperconnectivity may be due to normal ageing/physical activity was partially informed by the lack of correlation between longitudinal changes in functional connectivity and changes in subjective symptom burden. Subjective symptom reports have been documented to have reliability issues, particularly in the absence of a clinically diagnosed mTBI [64–66]. Moreover, symptom data was not available for the comparison group presented in this work. Fourth, a network-based voxel-wise analysis approach was applied to rs-fMRI data. While this is a common approach, it may lack the sensitivity to detect subtle cross-sectional differences in brain function. A more advanced technique like functional connectome analysis may be more sensitive to alterations in network efficiency related to HAE exposure. Fifth, this study focused on a single neuroimaging modality – rs-fMRI. Other modalities have demonstrated merit to study the impact of HAE exposure on the brain, particularly diffusion tensor imaging [29]. In a recently published study, our group observed notably different trends in white matter responses to HAE exposure in the same cohort described here [67]. These divergent trends further raise the complexity of the issue at hand. Multi-modal analytical approaches should be explored in future research to understand the overall neurological response to HAE exposure. Finally, our sample consists of only adolescent males participating in rugby union, limiting the generalizability of the findings to a wider population.

## Conclusion

Contrary to our hypotheses, no significant categorical differences in network-based functional connectivity were observed between collision sport and non-collision sport adolescent male athletes at any stage of the season. Similarly, functional connectivity at the end of the season did not differ between collision sport athletes stratified by high or low subclinical HAE exposure, based on the number of matches played, and the non-collision sport comparison group. Longitudinal analysis within the collision sport cohort revealed a pattern of hyperconnectivity in multiple networks over the season. However, longitudinal changes in functional connectivity were not associated with self-reported assessment of symptoms commonly associated with mTBI measured using the SCAT5. The longitudinal findings may represent compensatory adaptations to preserve neurological function during periods of HAE exposure. It is also plausible that these changes may be explained by normal ageing processes, effects of physical activity on developing brain networks, and/or motor learning. This finding highlights the complexity of interpreting changes in functional connectivity in adolescent athletes actively participating in collision sports.

## Data Availability

Data availability is limited due to data sovereignty considerations. Requests for access will be considered on a case by case basis.

## Acknowledgements

The authors acknowledge the participants, their whānau (family), Gisborne Boys High School, Poverty Bay Rugby Football Union, Mātai Ngā Mangai Māori, Turanga Health, and the wider Gisborne-Tairāwhiti community for their support and partnership toward this ongoing initiative. The authors would also like to acknowledge Dr Patrick McHugh and Professor Helen Danesh-Meyer for assisting with the initial organisation of this study. Thanks are given to the New Zealand eScience Infrastructure (NeSI) for high performance computing and consultancy services, we particularly thank Dr Maxime Rio. We acknowledge the assistance of Dr Olivia Harrison and Dr Sam Harrison in setting up the FSL analysis performed in this manuscript.

## References

1. Rice, S.G., Medical conditions affecting sports participation. Pediatrics, 2008. 121(4): p. 841–8.

2. World Rugby, Global rugby participation increasing ahead of Rugby World Cup 2023. 2023.

3. International Ice Hockey Federation, Survey of players.

4. National Football Foundation, Football by the numbers. 2023.

5. Kunz, M., 265 million playing football, in FIFA Magazine. 2007. p. 11-15.

6. Koutures, C.G., et al., Injuries in Youth Soccer. Pediatrics, 2010. 125(2): p. 410–414.

7. Bailey, R., Physical education and sport in schools: a review of benefits and outcomes. J Sch Health, 2006. 76(8): p. 397–401.

8. Merkel, D.L., Youth sport: positive and negative impact on young athletes. Open Access J Sports Med, 2013. 4: p. 151–60.

9. Nowinski, C.J., et al., Applying the Bradford Hill Criteria for Causation to Repetitive Head Impacts and Chronic Traumatic Encephalopathy. Front Neurol, 2022. 13: p. 938163.

10. Oldham, J.R., et al., *Neurocognitive functioning and symptoms across levels of collision and contact in male high school athletes.* Journal of Neurology, Neurosurgery & Psychiatry, 2022. 93(8): p. 828.

11. Hume, P.A., et al., A Comparison of Cognitive Function in Former Rugby Union Players Compared with Former Non-Contact-Sport Players and the Impact of Concussion History. Sports Med, 2017. 47(6): p. 1209–1220.

12. Bellomo, G., et al., A systematic review on the risk of neurodegenerative diseases and neurocognitive disorders in professional and varsity athletes. Neurological Sciences, 2022. 43(12): p. 6667–6691.

13. Ueda, P., et al., Neurodegenerative disease among male elite football (soccer) players in Sweden: a cohort study. The Lancet Public Health, 2023. 8(4): p. e256–e265.

14. Kuo, C., et al., On-Field Deployment and Validation for Wearable Devices. Annals of Biomedical Engineering, 2022. 50(11): p. 1372–1388.

15. Silverberg, N.D., et al., The American Congress of Rehabilitation Medicine Diagnostic Criteria for Mild Traumatic Brain Injury. Arch Phys Med Rehabil, 2023. 104(8): p. 1343–1355.

16. Pfister, T., et al., The incidence of concussion in youth sports: a systematic review and meta-analysis. Br J Sports Med, 2016. 50(5): p. 292–7.

17. Kerr, Z.Y., et al., Concussion Incidence and Trends in 20 High School Sports. Pediatrics, 2019. 144(5): p. e20192180.

18. Bussey, M.D., et al., Head Acceleration Events in Male Community Rugby Players: An Observational Cohort Study across Four Playing Grades, from Under-13 to Senior Men. Sports Med, 2024. 54(2): p. 517–530.

19. Holcomb, T.D., et al., Characterization of Head Acceleration Exposure During Youth Football Practice Drills. Journal of Applied Biomechanics, 2023. 39(3): p. 157–168.

20. Mez, J., et al., Duration of American Football Play and Chronic Traumatic Encephalopathy. Ann Neurol, 2020. 87(1): p. 116–131.

21. Adams, J.W., et al., Lewy Body Pathology and Chronic Traumatic Encephalopathy Associated With Contact Sports. J Neuropathol Exp Neurol, 2018. 77(9): p. 757–768.

22. McKee, A.C., et al., The spectrum of disease in chronic traumatic encephalopathy. Brain, 2013. 136(Pt 1): p. 43–64.

23. Bieniek, K.F., et al., Association between contact sports participation and chronic traumatic encephalopathy: a retrospective cohort study. Brain Pathology, 2020. 30(1): p. 63–74.

24. Fortington, L.V., et al., Epidemiological Principles in Claims of Causality: An Enquiry into Repetitive Head Impacts (RHI) and Chronic Traumatic Encephalopathy (CTE). Sports Medicine, 2024.

25. Mayer, A.R., et al., Radiologic common data elements rates in pediatric mild traumatic brain injury. Neurology, 2020. 94(3): p. e241–e253.

26. Bobholz, S.A., et al., Prospective study of the association between sport-related concussion and brain morphometry (3T-MRI) in collegiate athletes: Study from the NCAA-DoD CARE Consortium. British Journal of Sports Medicine, 2021. 55(3): p. 169–174.

27. Morelli, N., et al., Resting state functional connectivity responses post-mild traumatic brain injury: a systematic review. Brain Injury, 2021. 35(11): p. 1326–1337.

28. Lindsey, H.M., et al., Diffusion-Weighted Imaging in Mild Traumatic Brain Injury: A Systematic Review of the Literature. Neuropsychol Rev, 2021. 33(1): p. 42–121.

29. Koerte, I.K., et al., Diffusion Imaging of Sport-related Repetitive Head Impacts-A Systematic Review. Neuropsychol Rev, 2023. 33(1): p. 122–143.

30. 30. New Zealand Rugby, *New Zealand Rugby annual report 2023*. 2023.

31. Kawas, M.I., et al., Cognitive and Salience Network Connectivity Changes following a Single Season of Repetitive Head Impact Exposure in High School Football. AJNR Am J Neuroradiol, 2024. 45(8): p. 1116–1123.

32. Fitzgerald, B., et al., Longitudinal changes in resting state fMRI brain self-similarity of asymptomatic high school American football athletes. Sci Rep, 2024. 14(1): p. 1747.

33. DeSimone, J.C., et al., Mapping default mode connectivity alterations following a single season of subconcussive impact exposure in youth football. Hum Brain Mapp, 2021. 42(8): p. 2529–2545.

34. Murugesan, G., et al., Single Season Changes in Resting State Network Power and the Connectivity between Regions: Distinguish Head Impact Exposure Level in High School and Youth Football Players. Proc SPIE Int Soc Opt Eng, 2018. 10575.

35. Harris, P.A., et al., Research electronic data capture (REDCap)--a metadata-driven methodology and workflow process for providing translational research informatics support. J Biomed Inform, 2009. 42(2): p. 377–81.

36. Echemendia, R.J., et al., The Sport Concussion Assessment Tool 5th Edition (SCAT5): Background and rationale. Br J Sports Med, 2017. 51(11): p. 848–850.

37. Köster, J. and S. Rahmann, Snakemake—a scalable bioinformatics workflow engine. Bioinformatics, 2012. 28(19): p. 2520–2522.

38. Gorgolewski, K.J., et al., The brain imaging data structure, a format for organizing and describing outputs of neuroimaging experiments. Scientific Data, 2016. 3(1): p. 160044.

39. 39. Halchenko, Y.O., et al., *HeuDiConv — flexible DICOM conversion into structured directory layouts (v1.3.2).* Zenodo, 2024.

40. Esteban, O., et al., MRIQC: Advancing the automatic prediction of image quality in MRI from unseen sites. PLoS One, 2017. 12(9): p. e0184661.

41. Esteban, O., et al., fMRIPrep: a robust preprocessing pipeline for functional MRI. Nature Methods, 2019. 16(1): p. 111–116.

42. Varoquaux, G., et al., Multi-subject dictionary learning to segment an atlas of brain spontaneous activity. Inf Process Med Imaging, 2011. 22: p. 562–73.

43. Desikan, R.S., et al., An automated labeling system for subdividing the human cerebral cortex on MRI scans into gyral based regions of interest. Neuroimage, 2006. 31(3): p. 968–80.

44. Manning, K.Y., et al., Longitudinal changes of brain microstructure and function in nonconcussed female rugby players. Neurology, 2020. 95(4): p. e402–e412.

45. Sours, C., et al., Investigation of Multiple Frequency Ranges Using Discrete Wavelet Decomposition of Resting-State Functional Connectivity in Mild Traumatic Brain Injury Patients. Brain connectivity, 2015. 5(7): p. 442–450.

46. Dall’Acqua, P., et al., Functional and Structural Network Recovery after Mild Traumatic Brain Injury: A 1-Year Longitudinal Study. Frontiers in human neuroscience, 2017. 11: p. 280.

47. Palacios, E.M., et al., Resting-state functional connectivity alterations associated with six-month outcomes in mild traumatic brain injury. Journal of Neurotrauma, 2017. 34(8): p. 1546–1557.

48. Manning, K.Y., et al., Multiparametric MRI changes persist beyond recovery in concussed adolescent hockey players. Neurology, 2017. 89(21): p. 2157–2166.

49. D’Souza, M.M., et al., Alterations of connectivity patterns in functional brain networks in patients with mild traumatic brain injury: A longitudinal resting-state functional magnetic resonance imaging study. Neuroradiology Journal, 2020. 33(2): p. 186–197.

50. Fornito, A., A. Zalesky, and M. Breakspear, The connectomics of brain disorders. Nature Reviews Neuroscience, 2015. 16(3): p. 159–172.

51. van Duijvenvoorde, A.C.K., et al., A three-wave longitudinal study of subcortical-cortical resting-state connectivity in adolescence: Testing age– and puberty-related changes. Hum Brain Mapp, 2019. 40(13): p. 3769–3783.

52. Ploughman, M., Exercise is brain food: the effects of physical activity on cognitive function. Dev Neurorehabil, 2008. 11(3): p. 236–40.

53. Gomez-Pinilla, F. and C. Hillman, The influence of exercise on cognitive abilities. Compr Physiol, 2013. 3(1): p. 403–28.

54. Brooks, S.J., S.M. Parks, and C. Stamoulis, Widespread Positive Direct and Indirect Effects of Regular Physical Activity on the Developing Functional Connectome in Early Adolescence. Cerebral Cortex, 2021. 31(10): p. 4840–4852.

55. Li, J., et al., The alterations of functional brain networks and its relationship with sport decision-making and training duration in soccer players across different skill levels. Neurosci Lett, 2024. 831: p. 137788.

56. Sommer, M., et al., Timing Training in Female Soccer Players: Effects on Skilled Movement Performance and Brain Responses. Front Hum Neurosci, 2018. 12: p. 311.

57. Zhou, Q., et al., Long-term motor training enhances functional connectivity between semantic and motor regions in an effector-specific manner: evidence from elite female football athletes. Brain Struct Funct, 2024. 229(6): p. 1447–1459.

58. Knowles, S.B., S.W. Marshall, and K.M. Guskiewicz, Issues in estimating risks and rates in sports injury research. J Athl Train, 2006. 41(2): p. 207–15.

59. Anderson, E., et al., *Sport Structured Brain Trauma is Child Abuse.* Sport, Ethics and Philosophy, 2023: p. 1–21.

60. Richter, E.D. and R. Laster, The Precautionary Principle, epidemiology and the ethics of delay. Int J Occup Med Environ Health, 2004. 17(1): p. 9–16.

61. Mackay, D.F., et al., Neurodegenerative Disease Mortality among Former Professional Soccer Players. New England Journal of Medicine, 2019. 381(19): p. 1801–1808.

62. Eather, N., et al., The impact of sports participation on mental health and social outcomes in adults: a systematic review and the ‘Mental Health through Sport’ conceptual model. Systematic Reviews, 2023. 12(1): p. 102.

63. Strauss, S.B., et al., Framing potential for adverse effects of repetitive subconcussive impacts in soccer in the context of athlete and non-athlete controls. Brain Imaging Behav, 2021. 15(2): p. 882–895.

64. Voormolen, D.C., et al., Prevalence of post-concussion-like symptoms in the general population in Italy, The Netherlands and the United Kingdom. Brain Inj, 2019. 33(8): p. 1078–1086.

65. van der Vlegel, M., et al., Prevalence of Post-Concussion-Like Symptoms in the General Injury Population and the Association with Health-Related Quality of Life, Health Care Use, and Return to Work. Journal of clinical medicine, 2021. 10(4): p. 806.

66. Iverson, G.L., et al., Factors Associated With Concussion-like Symptom Reporting in High School Athletes. JAMA pediatrics, 2015. 169(12): p. 1132–1140.

67. Tayebi, M., et al., Characterizing the Effect of Repetitive Head Impact Exposure and mTBI on Adolescent Collision Sports Players’ Brain with Diffusion Magnetic Resonance Imaging. Journal of Neurotrauma, 2024.

